# Impact of preoperative thyroid status on BMI change after lifestyle intervention and bariatric surgery: Results from the OBESEPI cohort

**DOI:** 10.64898/2026.04.30.26352121

**Authors:** Claire Nominé-Criqui, Florence Bihain, Lola Bachelin, Nicolas Scheyer, Laurent Brunaud, David Meyre

## Abstract

**Background:** Obesity is a chronic multifactorial disease characterized by substantial interindividual variability in weight loss after lifestyle intervention and bariatric surgery. Thyroid hormones play a key role in energy homeostasis, but their influence on postoperative weight outcomes remains insufficiently studied.

**Objective:** To evaluate the association between preoperative thyroid status and changes in body mass index (BMI) after lifestyle intervention and bariatric surgery over a five-year follow-up.

**Methods:** We conducted a retrospective cohort study including adults with class II or III obesity enrolled in the Obésité Sévère et Épigénétique (OBESEPI) study. All participants underwent preoperative lifestyle intervention followed by bariatric surgery. Thyroid status was classified as euthyroid or hypothyroid based on clinical and biochemical criteria. BMI was assessed at baseline and at nine postoperative time points over five years.

**Results:** Among 435 included patients, 71 (16.8%) had hypothyroidism. Baseline BMI was similar between groups, whereas diabetes was more frequent in hypothyroid patients (52.1% vs 37.7%; p = 0.022). Hypothyroid patients had significantly higher BMI at 6–24 months after surgery, but differences were no longer significant beyond three years. BMI trajectories and magnitude of weight regain were comparable between groups. Higher preoperative TSH levels were independently associated with BMI regain (OR 1.32, 95% CI 1.00–1.72; p = 0.047). Higher baseline BMI, younger age, and female sex were also associated with greater BMI regain.

**Conclusions:** Hypothyroidism was associated with lower early postoperative weight loss but did not influence long-term BMI trajectories. Higher preoperative TSH levels were independently associated with BMI regain.

**KEYPOINTS:**

- Preoperative hypothyroidism is associated with reduced early weight loss during the first two years after bariatric surgery.
- Long-term BMI trajectories and weight regain patterns are similar between hypothyroid and euthyroid patients beyond three years of follow-up.
- Higher preoperative TSH levels independently predict BMI regain.
- Baseline BMI, younger age, and female sex remain key determinants of the magnitude of BMI regain after bariatric surgery.

## INTRODUCTION

According to the World Health Organization, 43% of adults globally were overweight or obese in 2022, representing a major public health challenge and a substantial economic burden (1,2). Despite the availability of multiple treatment strategies, including lifestyle modification, pharmacotherapy, and bariatric surgery, long-term weight management remains difficult, with considerable interindividual variability in treatment response (3). Bariatric surgery is currently the most effective and durable treatment for severe obesity (4,5). However, postoperative weight loss varies considerably between patients, and weight regain some time after surgery is frequently observed (3). Demographic, clinical, behavioral, genetic, and psychological factors may partly explain this variability, suggesting that other biological determinants may also play a role (6–8). Thyroid hormones are key regulators of energy balance through their effects on thermogenesis, resting metabolic rate, lipid metabolism, and appetite regulation (9). Hypothyroidism is associated with weight gain and reduced metabolic rate and affects approximately 1–2% of the general population (10,11). Evidence suggests a bidirectional relationship between obesity and thyroid function, with obesity associated with higher thyrotropin levels and increased risk of hypothyroidism (9,12), while weight loss may improve thyroid function. Emerging data indicate that thyroid dysfunction could influence postoperative outcomes and responsiveness to lifestyle interventions (13,14). However, the long-term impact of initial thyroid status (at the time of treatment initiation) on weight changes after bariatric surgery remains unclear and underreported. The aim of this study was to investigate the association between initial thyroid status and BMI changes during lifestyle intervention and over five years after bariatric surgery in a cohort of patients with severe obesity enrolled in the OBESEPI study.

## MATERIAL AND METHODS

### Study design and population

This retrospective cohort study included adults with class II or III obesity (BMI ≥35 kg/m^2^) enrolled in the Obésité Sévère et Épigénétique (OBESEPI) study conducted at Nancy University Hospital (ClinicalTrials.gov NCT02663388). Participants completed a structured lifestyle and behavioural intervention lasting approximately 12 months before bariatric surgery. Patients with hyperthyroidism, those who did not undergo surgery, or those with missing thyroid status data were excluded. The study was approved by the local ethics committee, and all participants provided written informed consent.

Thyroid status was determined using clinical history, medication records, and biochemical measurements obtained 1–6 months before surgery. Serum TSH and free T4 were measured using ultrasensitive ELISA assays. All patients with elevated TSH levels and/or those receiving thyroid hormone therapy were classified as hypothyroid. The remaining patients were classified as euthyroid.

BMI was recorded at baseline (initiation of care), on the day of surgery, and at 2, 6, 12, 18, 24, 36, 48, and 60 months after surgery. Postoperative weight evolution was evaluated using : longitudinal BMI values during follow-up, maximal BMI loss after surgery, BMI regain (increase >0.5 BMI units after nadir), magnitude of regain (BMImax ™ BMImin), time from BMI nadir to maximal regain, BMI trajectory patterns (L, U, D, Y curves) based on previously described classifications [5,7].

The primary objective was to assess the impact of thyroid status on postoperative BMI changes over five years. Secondary objectives included comparisons of metabolic characteristics, preoperative BMI change, weight regain, and magnitude of regain according to thyroid status.

### Statistical Analysis

Continuous variables were expressed as mean ± standard deviation or median (interquartile range) depending on distribution assessed using the Shapiro–Wilk test. Categorical variables were expressed as counts and percentages. Between-group comparisons were performed using Student’s t-test or Mann–Whitney U test for continuous variables and Chi-square or Fisher’s exact test for categorical variables. Multivariable logistic regression was used to identify factors associated with BMI regain (yes/no), magnitude of BMI regain (delta BMI), the first and second multivariate analysis was conducted on the complete cases and the third with imputation. Variables with p < 0.20 in univariate analysis were included in the multivariable model. Results were reported as odds ratios (OR) with 95% confidence intervals (CI). Model discrimination was assessed using the area under the receiver operating characteristic curve (AUC), and calibration was evaluated using the Hosmer–Lemeshow test. To account for missing data, multiple imputation by chained equations was performed, generating 20 imputed datasets. All analyses were conducted using R software (version 4.4.1). Statistical significance was defined as p < 0.05

## RESULTS

### Study population

Among the 454 patients initially enrolled in the cohort, 19 (4.2%) were excluded. **(Figure 1)**. After exclusion, the final dataset included 435 patients. Among those, a total of 71 patients (16.8%) were classified as hypothyroidism.

**Figure 1.**
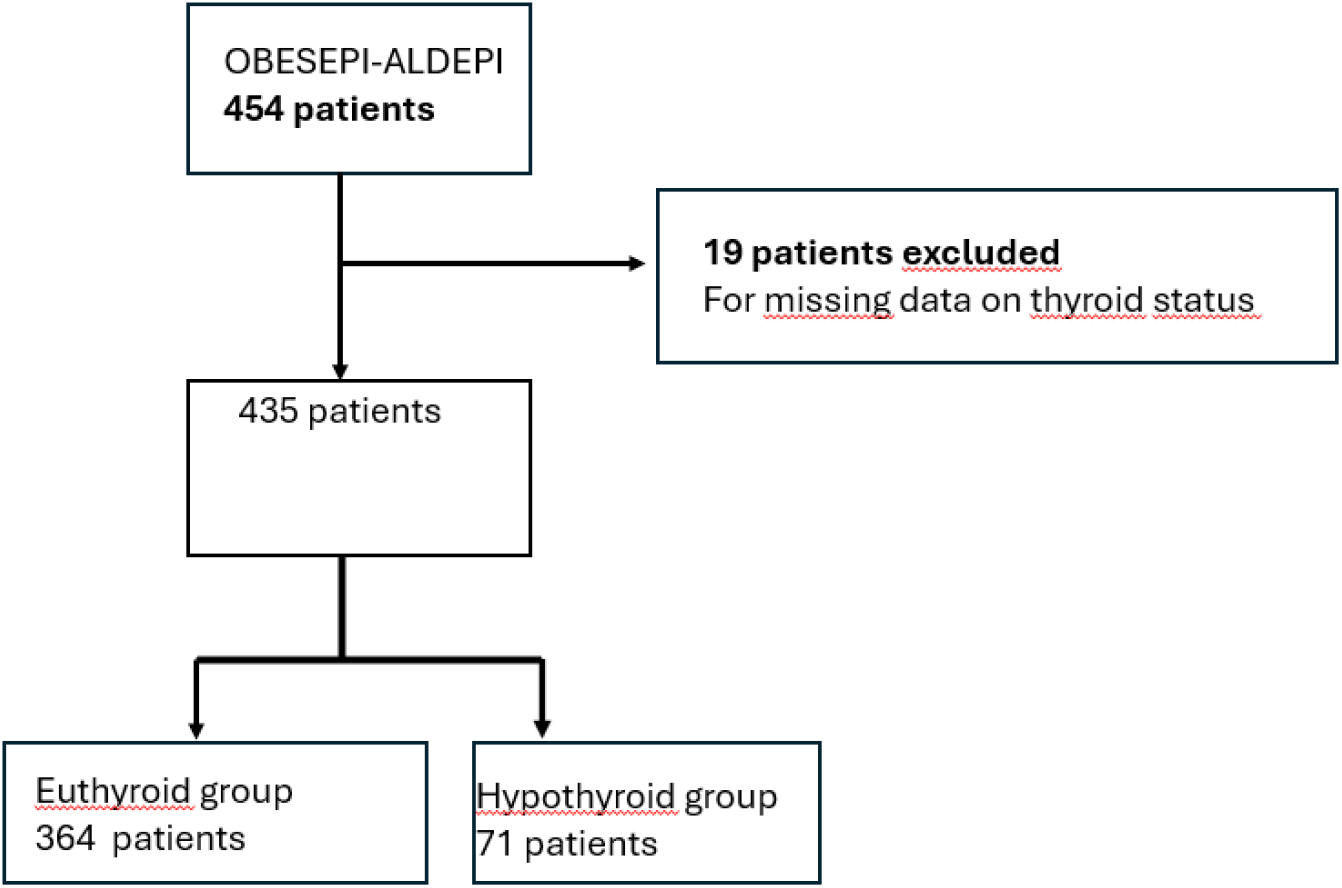
Flow-chart of the study

Patients classified as hypothyroid were more likely to be female (90.4% vs. 75.4%; p=0.0048) and older (47.41 + 11.57 vs. 42.72 + 11.4 yo; p=0.0015) than patients classified as euthyroid. These hypothyroid patients had higher TSH levels (3.154 + 2.249 vs. 2.099 + 0.808 mIU/L; p < 0.0001) and higher T4 levels (12.64 + 3.18 vs 10,96 + 1.99 pmol/L; p = 0.0011). Baseline weight and BMI did not differ significantly between hypothyroid and euthyroid patients. Hypertension and dyslipidaemia prevalence were similar between groups, whereas diabetes was more frequent among hypothyroid patients (52.06% vs 37.67%; p = 0.022) **(Table 1)**. The BMI variation between the initiation of care (baseline) and the day of surgery was similar between the two groups (1.98 in hypothyroid group vs 1.42 in euthyroid group; p=0.44). The number of patients with complete data during the follow-up is available in **Figure 2**.

**Table 1.**
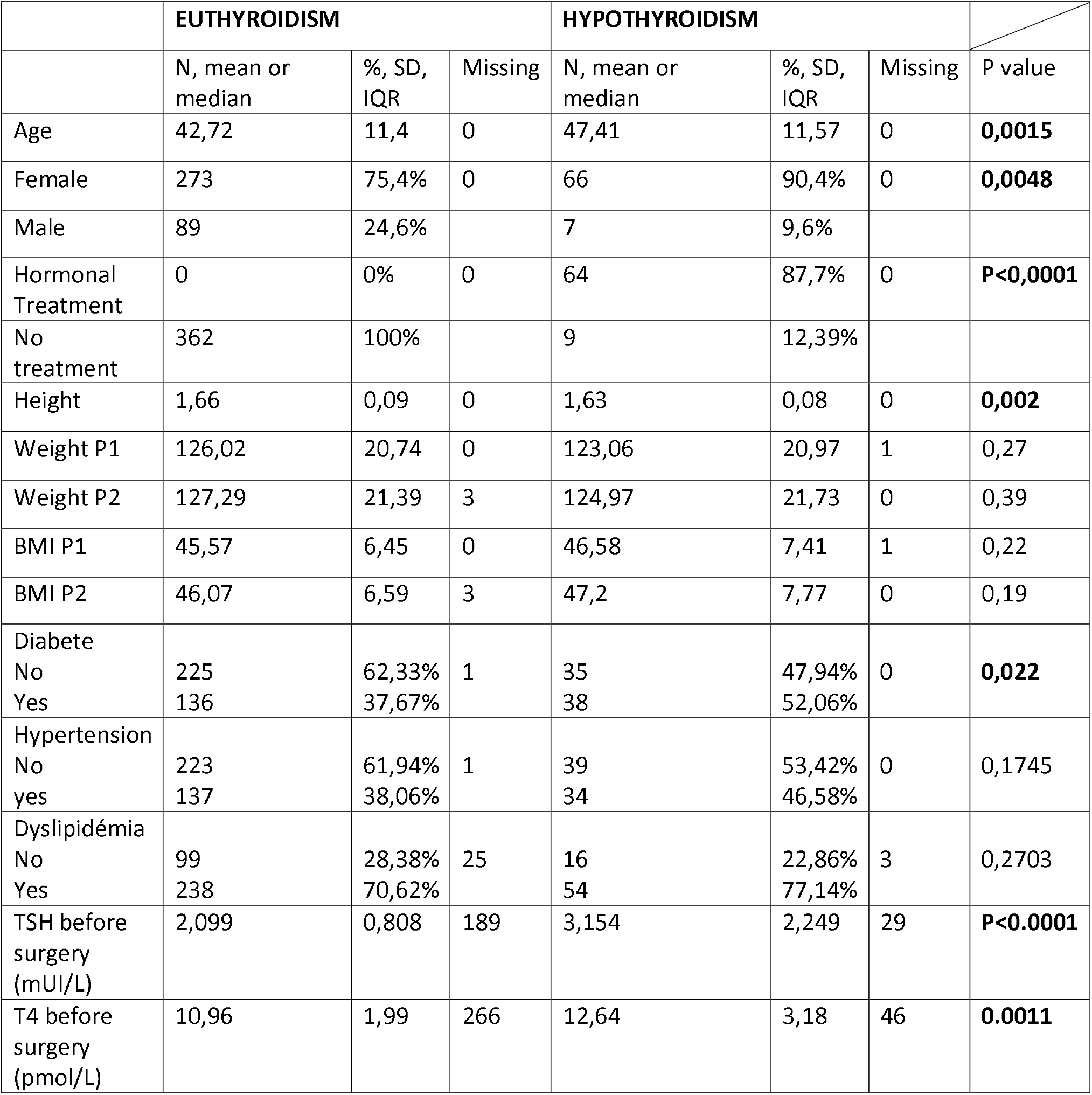
characteristics of the patients.

**Figure 2.**
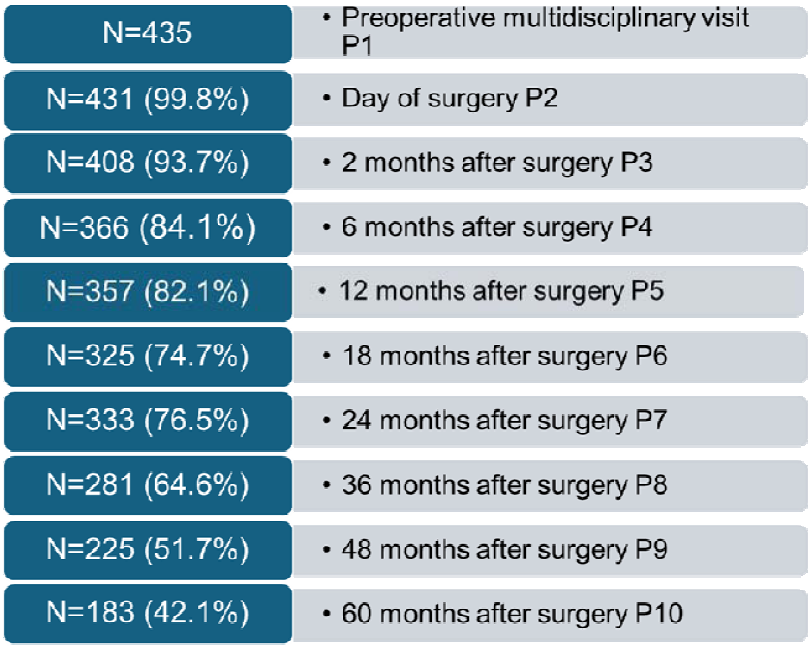
number of patients with complete data during the follow-up

### Weight loss after bariatric surgery

Participants underwent one of the following procedures: laparoscopic Roux-en-Y gastric bypass (91%-n=396), laparoscopic sleeve gastrectomy (8%, n=35), duodenal switch with or without Roux-en-Y (1.% ; n=4). Total weight loss differed significantly between groups at 6, 12, 18, and 24 months after surgery, with lower weight loss observed in patients classified as hypothyroid **(Table 2)**. From the third year after surgery, there was no longer any significant difference between the two groups in terms of weight loss. In a similar manner, BMI was significantly higher in hypothyroid patients at 6 months (36.99 vs 35.14; p = 0.0047), 12 months (33.64 vs 31.78; p = 0.0046), and 18 months (32.71 vs 30.63; p = 0.003) after surgery. No significant differences were observed beyond the third year after surgery.

**Table 2.** Changes in BMI after surgery.

|  | EUTHYROIDISM |  |  | HYPOTHYROIDISM |  |  |  |
| --- | --- | --- | --- | --- | --- | --- | --- |
|  | N, mean or median | % sd or IQR | missing | N, mean or median | % sd or IQR | missing | P Value |
| BMI P3(kg/m<sup>2</sup>) | 40.65 | 6.55 | 22 | 41.61 | 7.73 | 4 | 0.2904 |
| BMI P4(kg/m<sup>2</sup>) | 35.14 | 6.42 | 60 | 36.99 | 8.01 | 8 | **0.00472** |
| BMI P5 (kg/m<sup>2</sup>) | 31.78 | 6.19 | 67 | 33.64 | 7.61 | 10 | **0.00466** |
| BMI P6 (kg/m<sup>2</sup>) | 30.63 | 6.08 | 97 | 32.71 | 8.22 | 12 | **0.00266** |
| BMI P7 (kg/m<sup>2</sup>) | 30.34 | 6.43 | 88 | 32.96 | 8.39 | 13 | 0.081 |
| BMI P8 (kg/m<sup>2</sup>) | 30.96 | 6.56 | 132 | 32.93 | 6.87 | 21 | 0.0576 |
| BMI P9 (kg/m<sup>2</sup>) | 31.21 | 6.48 | 179 | 32.37 | 7.35 | 34 | 0.3312 |
| BMI P10 (kg/m<sup>2</sup>) | 31.51 | 6.48 | 211 | 32.35 | 6.48 | 40 | 0.5145 |
| Delta BMI P1P2 | 1.42 | 5.8 | 3 | 1.98 | 5.26 | 1 | 0.440 |
| Delta BMI P1P3 | -13.63 | 8.78 | 21 | -13.04 | 8.4 | 6 | 0.612 |
| Delta BMI P1P4 | -28.67 | 12.07 | 59 | -25.43 | 9.71 | 10 | **0.0462** |
| Delta BMI P1P5 | -37.65 | 13.17 | 66 | -33.59 | 11.06 | 12 | **0.024** |
| Delta BMI P1P6 | -41.17 | 14.43 | 97 | -36.63 | 12.62 | 13 | **0.025** |
| Delta BMI P1P7 | -41.81 | 14.84 | 88 | -37.35 | 11.53 | 14 | **0.036** |
| Delta BMI P1P8 | -39.76 | 15.02 | 132 | -35.41 | 11.88 | 22 | 0.054 |
| Delta BMI P1P9 | -38.75 | 15.22 | 175 | -36.73 | 13.12 | 35 | 0.447 |
| Delta BMI P1P10 | -38.76 | 15.43 | 211 | -36.37 | 10.75 | 41 | 0.405 |
| BMI Regain |  |  |  |  |  |  |  |
| Yes | 246 | 75,93% | 38 | 52 | 76,47% | 5 | 0,92 |
| no | 78 | 24,07% |  | 16 | 23,53% |  |  |
| Delta BMI regain | 3,849 | 2,591 | 116 | 3,544 | 2,66 | 21 | 0,4427 |
| Time between min BMI/max BMI (in months) | 37,163 | 25,345 | 116 | 34,846 | 26,356 | 52 | 0,0552 |
| Time of BMI min (in months) | 20,54 | 9,91 | 116 | 21,923 | 10,55 | 21 | 0,3726 |

### Weight regain and BMI trajectories

After the second year after surgery, the mean BMI increased in the euthyroid group, while it continued to decrease in the hypothyroid group patients. The distribution of BMI trajectory profiles (L, U, D, Y curves) did not differ significantly between hypothyroid and euthyroid patients when grouped as weight regain versus no regain (76.5% vs 75.9%; p = 0.92).

Among patients with weight regain, the magnitude of regain was similar between groups (3.54 vs 3.85 BMI units; p = 0.445), as was the time to reach minimum postoperative weight (21.92 vs 20.54 months; p = 0.37). Between two and three years postoperatively, BMI changes were comparable between groups (0.634 vs 0.757; p = 0.69). However, a lower proportion of hypothyroid patients began to regain weight during this period compared with euthyroid patients (60% vs 74.4% p = 0.041) **(Table 3)**.

**Table 3.** weight variation, BMI variation and regain between 2 and 3 years after surgery.

|  | EUTHYROIDISM |  |  | HYPOTHYROIDISM |  |  | P Value |
| --- | --- | --- | --- | --- | --- | --- | --- |
|  | N, mean or median | % sd or IQR | missing | N, mean or median | % sd or IQR | missing |  |
| BMI between 2 and 3 years after surgery | 0,757 | 1,983 | 143 | 0,634 | 1,888 | 23 | 0,6924 |
| No regain between 2 and 3 years after surgery | 56 | 25.57% | 143 | 20 | 40% | 23 | **0,0409** |
| Regain between 2 and 3 years after surgery | 163 | 74.43% | 143 | 30 | 60% | 23 | **0,0409** |

### Factors associated with BMI regain (yes/no)

A total of 437 patients were included in the logistic regression analysis, of whom 143 had complete data. In complete-case multivariable analysis, no variable was independently associated with BMI regain **(Table 4)**. The model demonstrated modest discrimination (AUC = 0.68). **(Figure 3)**

**Table 4.** Factors associated with BMI regain (yes/no) in Multivariate analysis with OR.

| Variables | OR (CI 95%) | p |
| --- | --- | --- |
| Hormonal Treatment | 0,7 (0,03-8,07) | 0,79 |
| TSH (mUI/L) | 1,32(0,99_1,92) | 0,125 |
| Age | 1,03(0,93-1,07) | 0,19 |
| Sex | 1,09 (0,41-2,78) | 0,86 |
| Dyslipidemia | 0,37(0,11-1,08) | 0,08 |
| Diabete | 1,36(0,58-3,29) | 0,48 |
| Hypertension | 0,85(0,35-2,11) | 0,73 |
| Surgery (By-pass) | 0,54(0,12-2,6) | 0,43 |
| BMIP1 | 1,02(0,95-1,09) | 0,615 |

**Figure 3.**
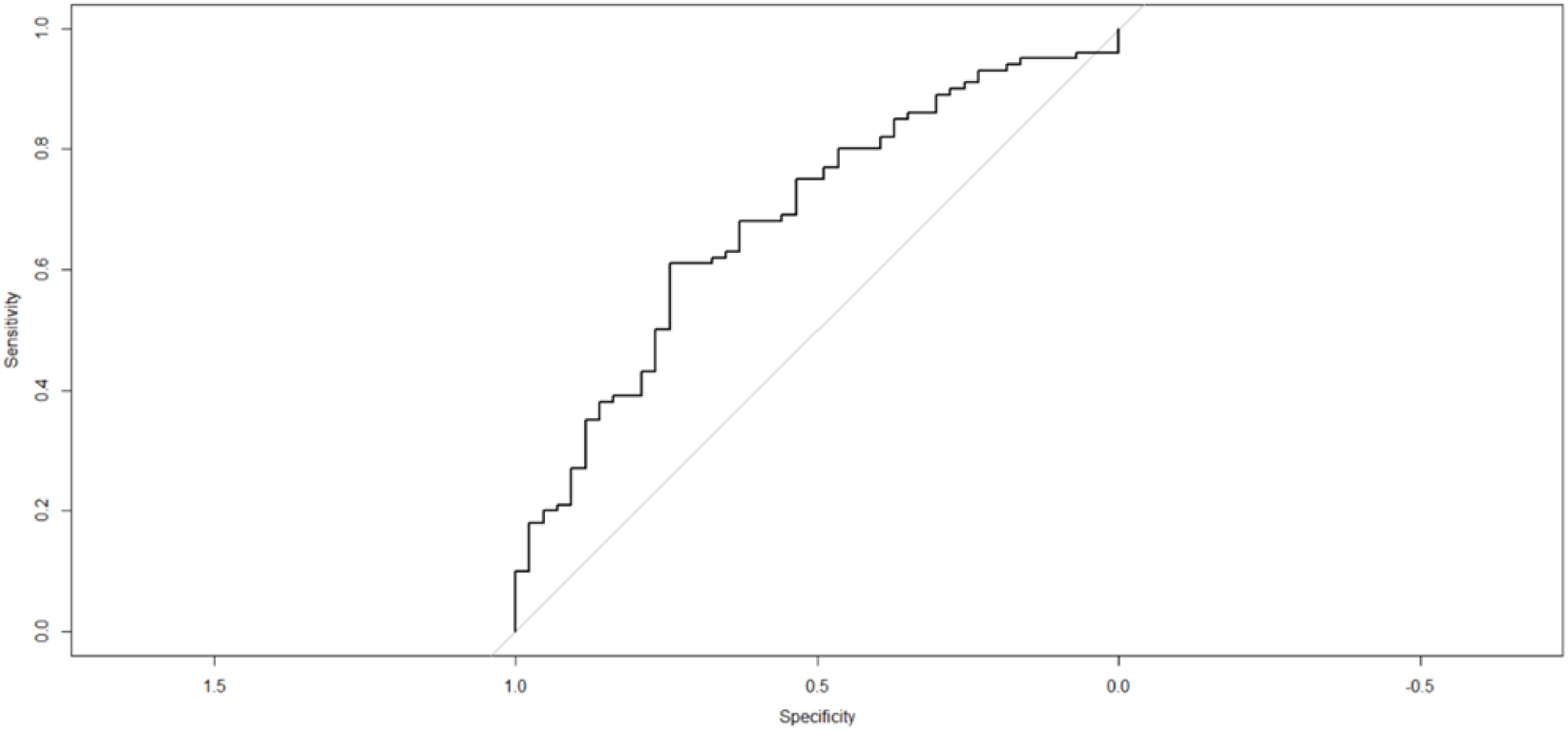
ROC Curve of the multivariate model.The multivariable model showed modest discrimination with an area under the receiver operating characteristic curve (AUC) of 0.68.

After multiple imputation, higher TSH levels were independently associated with increased odds of BMI regain (OR 1.32, 95% CI 1.00–1.72; p = 0.047). No other variables were significantly associated with BMI regain **(Table 5)**.

**Table 5.** Factors associated with BMI regain (yes/no) multivariate analysis with imputation.

| Variable | OR | 95%CI | P value |
| --- | --- | --- | --- |
| TSH (mIU/L) | 1.32 | 1-1.72 | **0.047** |
| Levothyroxine treatment | 0.63 | 1.1-3.88 | 0.619 |
| Age(years) | 1.02 | 0.99-1.04 | 0.231 |
| Female sex | 1.08 | 0.58-2.02 | 0.802 |
| Dyslipidemia | 0.68 | 0.38-1.22 | 0.197 |
| Diabetes | 1.19 | 0.68-2.1 | 0.545 |
| Hypertension | 1.03 | 1.03-0.58 | 0.920 |
| BMI at surgery (P1) | 0.98 | 0.94-1.03 | 0.418 |

### Factors associated with delta BMI regain

In multivariable linear regression, higher baseline BMI (BMI P1 at initiation of care (baseline)) was strongly associated with greater BMI regain (β = 0.37; p < 0.001). Older age was associated with lower regain (β = ™0.07; p = 0.005), and female sex was independently associated with regain magnitude (β = 1.82; p = 0.015). TSH levels and dyslipidaemia were not significantly associated with BMI regain magnitude. The model explained approximately 28% of the variance (adjusted R^2^ = 0.27) **(Table 6)**

**Table 6.** Multivariable Linear regression analysis of factors associated with delta BMI max.

| **Variable** | **B coefficient (95%CI)** | **p-value** |
| --- | --- | --- |
| TSH | -0.17(-0.62-0.29) | 0.478 |
| Age | -0.07(-0.12- -0.02) | 0.005 |
| BMIP1 | 0.37(0.27-0.46) | <0.001 |
| Dyslipidemia | -0.72(-2.08-0.64) | 0.297 |
| Sex (female) | 1.82(0.36 to 3.28) | 0.015 |

*B coefficients represent change in delta BMI max per unit increase in predictor. Analysis performed on complete cases*

## DISCUSSION

This study evaluated the impact of thyroid status on long-term weight changes after bariatric surgery. It showed that patients classified as hypothyroid had an initial delay in weight loss compared to patients classified as euthyroid, but that this difference disappeared approximately two to three years after surgery. After the second year following surgery, the mean BMI increased in the euthyroid group, while it continued to decrease in the hypothyroid group patients. These results suggest that thyroid function may modulate early postoperative metabolic adaptation, but that its influence diminishes over time. To our knowledge, this is the first study to evaluate weight changes according to preoperative thyroid status with a five-year follow-up after bariatric surgery.

It is considered that the incidence of hypothyroidism is between 1.2 to 5% of subjects in the general population (15). In a meta-analysis, Song et al. reported that obesity nearly doubles the overall risk of hypothyroidism (relative risk 1.86) and more than triples the risk of overt hypothyroidism (relative risk 3.21) compared with normal weight population (12). This study confirmed these findings, with a prevalence of hypothyroidism around 17%, which is significantly higher than in the general population (16).

Most prior studies comparing weight loss outcomes between hypothyroid and euthyroid patients have reported no significant differences after bariatric surgery. Szomstein et al. (17) and Raftopoulos et al. (18) reported comparable weight loss outcomes during the first postoperative year. However, these studies were limited by a relatively short follow-up period, which may have prevented the identification of a significant difference in postoperative weight with a longer follow-up. The data from this study were more closely aligned with those of Remmel et al. (19), which reported a reduction in the percentage of excess weight loss during the first postoperative year in hypothyroid patients.

Studies focusing solely on hypothyroid populations consistently demonstrated substantial weight loss following bariatric surgery, with total weight loss ranging from approximately 25% at six months to over 35% at one year (20–22). These results confirmed that hypothyroid status does not represent a major barrier for achieving clinically meaningful weight loss (delta BMI at 5 years : -36.37 vs -38.76; p=0.405), supporting the overall effectiveness of bariatric surgery even in this subgroup of patients. Weight loss was comparable between the two groups at 5 years However, this study findings suggested that hypothyroid patients should not be expected to follow identical early weight trajectories as euthyroid patients, but that long-term outcomes were likely comparable. The identification of a potential tipping point between two and three years after surgery also underscores the importance of continued endocrine and nutritional monitoring beyond the first postoperative year.

The association between thyroid dysfunction and systemic inflammation provides a plausible biological framework for interpreting this study data. Elevated inflammatory markers observed in hypothyroidism may improve following bariatric surgery and correlate with reductions in TSH levels independent of BMI changes (22,23). This supports the hypothesis that thyroid alterations in obese patients may represent an adaptive response to chronic inflammation rather than a primary endocrine disorder (24,25). The convergence of weight trajectories after two years observed in this study could reflect the improvement of inflammatory and neuroendocrine adaptations that initially blunt metabolic efficiency in hypothyroid patients before bariatric surgery.

In multivariate analysis, we observed that younger age and female sex were significantly associated with BMI regain. This finding may be partly explained by the imbalance in the male–female ratio in our cohort, with men representing only 22% of participants, which may have limited the statistical power to detect an effect of male sex on weight trajectories. In addition, some women may have experienced pregnancy during the follow-up period. Previous studies have shown that pregnancy could temporarily modify post-surgical weight-loss trajectories and contribute to gestational weight gain and postpartum weight retention (27–29) . We also observed that younger age was associated with weight regain. This is consistent with previous studies reporting that while younger patients generally achieved greater initial weight loss (30), they may also experience greater long-term weight regain compared with older patients (31,32).

In the multivariate analysis with imputation, we observed that higher TSH level was associated with regain status. Several mechanisms may explain this finding. First, obesity has been associated with moderately elevated TSH levels, possibly reflecting a functional adaptation of the hypothalamic–pituitary–thyroid axis to increased adiposity (33). In addition, previous studies have shown that thyroid hormone replacement frequently requires dose adjustment during postoperative follow-up, particularly during periods of rapid weight change (34,35). Consequently, higher TSH levels may also reflect difficulties in optimizing thyroid hormone replacement therapy during postoperative follow-up. Taken together, these findings suggested that TSH may act both as a marker of adiposity and as a potential indicator of suboptimal thyroid hormone replacement in patients experiencing unfavorable weight trajectories after bariatric surgery

In this cohort, interpreting the relationship between TSH levels and postoperative weight changes was complicated by the heterogeneity of the causes of hypothyroidism. Some patients were classified as hypothyroid after undergoing total thyroidectomy, while others had autoimmune thyroiditis. These conditions may have different effects on weight changes following bariatric surgery. Obesity has been associated with autoimmune thyroid disorders, and weight loss has been reported to reduce thyroid autoimmunity and normalize thyroid function parameters in some patients (36). In contrast, patients after total thyroidectomy require lifelong thyroid hormone replacement, and bariatric surgery may alter levothyroxine requirements through postoperative changes in body weight, body composition, and gastrointestinal absorption (34). Consequently, the association observed in this study between TSH levels and weight regain may in fact reflect different underlying mechanisms depending on the causes of hypothyroidism.

This study has several limitations. First, this study was observational and retrospective. Behavioral factors such as diet, physical activity, and psychological variables were not captured and may have influenced postoperative weight trajectories. In addition, lack of information regarding the cause of hypothyroidism prevented subgroup analyses. Lastly, this study suffers from a significant amount of missing data.

## CONCLUSION

In conclusion, preoperative hypothyroid status was associated with a modest delay in early postoperative weight loss after bariatric surgery. However, BMI trajectories converged within two to three years, suggesting that thyroid status may influence early metabolic adaptation rather than long-term weight outcomes. Future prospective studies incorporating comprehensive hormonal profiling are needed to clarify the causal role of thyroid function in long-term weight regulation after bariatric surgery.

## Data Availability

All data produced in the present study are available upon reasonable request to the authors

## REFERENCES

1. Fontbonne A, Currie A, Tounian P, Picot MC, Foulatier O, Nedelcu M, et al. Prevalence of Overweight and Obesity in France: The 2020 Obepi-Roche Study by the « Ligue Contre l’Obésité ». J Clin Med. 25 janv 2023;12(3):925.

2. Okunogbe A, Nugent R, Spencer G, Powis J, Ralston J, Wilding J. Economic impacts of overweight and obesity: current and future estimates for 161 countries. BMJ Glob Health. sept 2022;7(9):e009773.

3. Perdomo CM, Cohen RV, Sumithran P, Clément K, Frühbeck G. Contemporary medical, device, and surgical therapies for obesity in adults. Lancet Lond Engl. 1 avr 2023;401(10382):1116⍰30.

4. O’Brien PE, Hindle A, Brennan L, Skinner S, Burton P, Smith A, et al. Long-Term Outcomes After Bariatric Surgery: a Systematic Review and Meta-analysis of Weight Loss at 10 or More Years for All Bariatric Procedures and a Single-Centre Review of 20-Year Outcomes After Adjustable Gastric Banding. Obes Surg. janv 2019;29(1):3⍰14.

5. Karlsson J, Taft C, Rydén A, Sjöström L, Sullivan M. Ten-year trends in health-related quality of life after surgical and conventional treatment for severe obesity: the SOS intervention study. Int J Obes 2005. août 2007;31(8):1248⍰61.

6. McGarrity LA, Terrill AL, Martinez PL, Ibele AR, Morrow EH, Volckmann ET, et al. The Role of Resilience in Psychological Health Among Bariatric Surgery Patients. Obes Surg. mars 2022;32(3):792⍰800.

7. Sharma T, Morassut RE, Langlois C, Meyre D. Body mass index trajectories and their predictors in undergraduate students from Canada: Results from the GENEiUS study. J Am Coll Health J ACH. oct 2024;72(7):2147⍰55.

8. Blüher M. Obesity: global epidemiology and pathogenesis. Nat Rev Endocrinol. mai 2019;15(5):288⍰98.

9. Walczak K, Sieminska L. Obesity and Thyroid Axis. Int J Environ Res Public Health. 7 sept 2021;18(18):9434.

10. Wilson SA, Stem LA, Bruehlman RD. Hypothyroidism: Diagnosis and Treatment. Am Fam Physician. 15 mai 2021;103(10):605⍰13.

11. Vanderpump MPJ. The epidemiology of thyroid disease. Br Med Bull. 2011;99:39⍰51.

12. Song RH, Wang B, Yao QM, Li Q, Jia X, Zhang JA. The Impact of Obesity on Thyroid Autoimmunity and Dysfunction: A Systematic Review and Meta-Analysis. Front Immunol. 2019;10:2349.

13. Meneghini V, de Lima Beltrão FE, Golovko G, Watson RK, Samreen S, Ettleson MD, et al. Adverse outcomes in patients with hypothyroidism undergoing bariatric surgery: A retrospective study using TriNetX. J Clin Endocrinol Metab. 16 sept 2025;dgaf519.

14. Croce L, Pallavicini C, Crotti S, Coperchini F, Minnelli L, Magri F, et al. Basal and longitudinal changes in serum levels of TSH in morbid obese patients experiencing failure or success of dietary treatment. Eat Weight Disord EWD. août 2021;26(6):1949⍰55.

15. Vanderpump MP, Tunbridge WM, French JM, Appleton D, Bates D, Clark F, et al. The incidence of thyroid disorders in the community: a twenty-year follow-up of the Whickham Survey. Clin Endocrinol (Oxf). juill 1995;43(1):55⍰68.

16. Vanderpump MPJ. The epidemiology of thyroid disease. Br Med Bull. 1 sept 2011;99(1):39⍰51.

17. Szomstein S, Avital S, Brasesco O, Mehran A, Cabral JM, Rosenthal R. Laparoscopic gastric bypass in patients on thyroid replacement therapy for subnormal thyroid function-prevalence and short-term outcome. Obes Surg. janv 2004;14(1):95⍰7.

18. Raftopoulos Y, Gagné DJ, Papasavas P, Hayetian F, Maurer J, Bononi P, et al. Improvement of hypothyroidism after laparoscopic Roux-en-Y gastric bypass for morbid obesity. Obes Surg. avr 2004;14(4):509⍰13.

19. Remmel S, Noom M, Sandstrom R, Mhaskar R, Diab ARF, Sujka JA, et al. Preoperative comorbidities as a predictor of EBWL after bariatric surgery: a retrospective cohort study. Surg Endosc. mai 2024;38(5):2770⍰6.

20. Misra S, S C, S SK, S P, X L JL, P PR. Does Bariatric Surgery Impact Hypothyroidism and Vice Versa? Results of a Prospective Comparative Study. Obes Surg. oct 2025;35(10):4142⍰50.

21. Janssen IMC, Homan J, Schijns W, Betzel B, Aarts EO, Berends FJ, et al. Subclinical hypothyroidism and its relation to obesity in patients before and after Roux-en-Y gastric bypass. Surg Obes Relat Dis Off J Am Soc Bariatr Surg. 2015;11(6):1257⍰63.

22. Zhu C, Gao J, Mei F, Lu L, Zhou D, Qu S. Reduction in Thyroid-Stimulating Hormone Correlated with Improved Inflammation Markers in Chinese Patients with Morbid Obesity Undergoing Laparoscopic Sleeve Gastrectomy. Obes Surg. déc 2019;29(12):3954⍰65.

23. Shi C, Zhu L, Chen X, Gu N, Chen L, Zhu L, et al. IL-6 and TNF-α induced obesity-related inflammatory response through transcriptional regulation of miR-146b. J Interferon Cytokine Res Off J Int Soc Interferon Cytokine Res. mai 2014;34(5):342⍰8.

24. Matarese G. The link between obesity and autoimmunity. Science. 31 mars 2023;379(6639):1298⍰300.

25. Ruiz-Tovar J, Boix E, Galindo I, Zubiaga L, Diez M, Arroyo A, et al. Evolution of subclinical hypothyroidism and its relation with glucose and triglycerides levels in morbidly obese patients after undergoing sleeve gastrectomy as bariatric procedure. Obes Surg. mai 2014;24(5):791⍰5.

26. Nannipieri M, Cecchetti F, Anselmino M, Camastra S, Niccolini P, Lamacchia M, et al. Expression of thyrotropin and thyroid hormone receptors in adipose tissue of patients with morbid obesity and/or type 2 diabetes: effects of weight loss. Int J Obes. sept 2009;33(9):1001⍰6.

27. Ceulemans D, De Mulder P, Lebbe B, Coppens M, De Becker B, Dillemans B, et al. Gestational weight gain and postpartum weight retention after bariatric surgery: data from a prospective cohort study. Surg Obes Relat Dis Off J Am Soc Bariatr Surg. avr 2021;17(4):659⍰66.

28. Barajas-Gamboa JS, Khan MSI, Dang JT, Romero-Velez G, Diaz Del Gobbo G, Abdallah M, et al. The Effects of Post-Surgical Pregnancy on Weight Loss Trajectories after Bariatric Surgery: Are Initial Weight and Age Prognostic Factors? J Clin Med. 23 févr 2024;13(5):1264.

29. Xu H, Holowko N, Näslund I, Ottosson J, Arkema EV, Neovius M, et al. Pregnancy Weight Gain After Gastric Bypass or Sleeve Gastrectomy. JAMA Netw Open. 5 déc 2023;6(12):e2346228.

30. Contreras JE, Santander C, Court I, Bravo J. Correlation between age and weight loss after bariatric surgery. Obes Surg. août 2013;23(8):1286⍰9.

31. Livhits M, Mercado C, Yermilov I, Parikh JA, Dutson E, Mehran A, et al. Preoperative predictors of weight loss following bariatric surgery: systematic review. Obes Surg. janv 2012;22(1):70⍰89.

32. Monaco-Ferreira DV, Leandro-Merhi VA. Weight Regain 10 Years After Roux-en-Y Gastric Bypass. Obes Surg. mai 2017;27(5):1137⍰44.

33. Bétry C, Challan-Belval MA, Bernard A, Charrié A, Drai J, Laville M, et al. Increased TSH in obesity: Evidence for a BMI-independent association with leptin. Diabetes Metab. juin 2015;41(3):248⍰51.

34. Gadiraju S, Lee CJ, Cooper DS. Levothyroxine Dosing Following Bariatric Surgery. Obes Surg. oct 2016;26(10):2538⍰42.

35. Richou M, Gilly O, Taillard V, Paul De Brauwere D, Donici I, Guedj AM. Levothyroxine dose adjustment in hypothyroid patients following gastric sleeve surgery. Ann Endocrinol. oct 2020;81(5):500⍰6.

36. Bétry C, Challan-Belval MA, Bernard A, Charrié A, Drai J, Laville M, et al. Increased TSH in obesity: Evidence for a BMI-independent association with leptin. Diabetes Metab. juin 2015;41(3):248⍰51.

